# A Comprehensive Public Health Evaluation of Lockdown as a Non-pharmaceutical Intervention on COVID-19 Spread in India: National Trends Masking State Level Variations

**DOI:** 10.1101/2020.05.25.20113043

**Authors:** Deepankar Basu, Maxwell Salvatore, Debashree Ray, Mike Kleinsasser, Soumik Purkayastha, Rupam Bhattacharyya, Bhramar Mukherjee

**Author notes:** Corresponding author. <u>Address</u> - Department of Biostatistics, School of Public Health, University of Michigan, 1420 Washington Heights, Ann Arbor, MI 48109-2029, USA. <u>Telephone</u> –. <u>Email</u> –.

## Abstract

**Introduction:** India has been under four phases of a national lockdown from March 25 to May 31 in response to the COVID-19 pandemic. Unmasking the state-wise variation in the effect of the nationwide lockdown on the progression of the pandemic could inform dynamic policy interventions towards containment and mitigation.

**Methods:** Using data on confirmed COVID-19 cases across 20 states that accounted for more than 99% of the cumulative case counts in India till May 31, 2020, we illustrate the masking of state-level trends and highlight the variations across states by presenting evaluative evidence on some aspects of the COVID-19 outbreak: case-fatality rates, doubling times of cases, effective reproduction numbers, and the scale of testing.

**Results:** The estimated effective reproduction number R for India was 3.36 (95% confidence interval (CI): [3.03, 3.71]) on March 24, whereas the average of estimates from May 25 - May 31 stands at 1.27 (95% CI: [1.26, 1.28]). Similarly, the estimated doubling time across India was at 3.56 days on March 24, and the past 7-day average for the same on May 31 is 14.37 days. The average daily number of tests have increased from 1,717 (March 19-25) to 131,772 (May 25-31) with an estimated testing shortfall of 4.58 million tests nationally by May 31. However, various states exhibit substantial departures from these national patterns.

**Conclusions:** Patterns of change over lockdown periods indicate the lockdown has been effective in slowing the spread of the virus nationally. The COVID-19 outbreak in India displays large state-level variations and identifying these variations can help in both understanding the dynamics of the pandemic and formulating effective public health interventions. Our framework offers a holistic assessment of the pandemic across Indian states and union territories along with a set of interactive visualization tools that are daily updated at covind19.org.

## INTRODUCTION

Coronavirus disease of 2019 (COVID-19) is an infectious disease caused by severe acute respiratory syndrome coronavirus 2 (SARS-CoV-2).^1^ First identified in December 2019 in Wuhan, China, it has since spread globally, resulting in an ongoing pandemic.^2^ As of June 9, 2020, more than 7 million cases have been reported across 188 countries and territories, resulting in more than 405,000 deaths. India, a democracy of 1.35 billion with a high population density and fragile healthcare infrastructure is one of the global epicenters for this pandemic. In order to slow the spread of this pandemic, India implemented a strict nation-wide lockdown from March 25 until May 31, 2020^3^, after which phased lockdown for containment zones is in effect until June 30, 2020^4^. As of June 11, the number of total confirmed cases in India has crossed 298,000, of whom 8,501 have died and 146,972 have recovered, placing India at a worldwide rank of 4 in terms of total confirmed cases.^5^ The number of new cases in India are not on the decline even after nine weeks of national lockdown.

In light of tremendous public health interest, numerous data repositories, along with statistical models, are being developed with the aim of studying the effect of COVID-19 non-medical interventions. The focus of modeling is shifting from forecasting to evaluation of the effect of various interventions on the spread of the virus.^6^ As of June 9, 4,880 COVID-19 SARS-CoV-2 preprints have been uploaded to medRxiv and bioRxiv, of which at least 30 focus on analyzing the efficacy of the non-medical interventions implemented by the Indian government.

Ray and colleagues studied the short- and long-term impact of the initial lockdown on the total number of cases in India using standard epidemiological forecasting models, and concluded that the lockdown stood a good chance of reducing the total number of cases in India in the short term.^7^ Looking at several metrics, Mitra and colleagues suggested that curtailment strategies employed by the Indian government seem to have been effective in controlling the spread of the pandemic in India.^8^ Ghosh and colleagues investigated the spread of the virus and subsequent impact of preventive measures on the same at a state-level in India and noted that the lockdown has had differential effects on daily infection rates for various states in India.^9^ Jakhar and colleagues modeled data released by the Indian Ministry of Health and Family Welfare using the classical SIR (Susceptible-Infected-Recovered) model and calculated the basic reproduction number (R_0_) for India as a whole, along with state-specific values of the same.^10^ Similarly, Gupta and colleagues estimated key epidemiological parameters and evaluated the effect of control measures on the COVID-19 epidemic in India and its states using a dynamic compartment-based SEIR-QDPA modeling approach, reiterating that state-specific R_0_ values exhibit high variability with respect to the national value of R_0_.^11^ However, much is left to be done now that the nation-wide lockdown has ended and a targeted lockdown phase is ongoing. All epidemiological projections suggest that current gains may be reversed rapidly if air travel and social mixing resumes. For the time being, the general guideline is to re-open the country in a phased manner.^12^ The need of the hour is to study and analyze infection, recovery and fatality trends at a more granular level using multiple measures of assessing epidemic dynamics to ensure the formulation of targeted and customized interventions aimed at containment and mitigation.

In this paper, we consider an ensemble of metrics including case and death counts, case-fatality rates, effective basic reproduction numbers, doubling times, and assessment of testing shortfall across states for a deeper and policy-relevant understanding of the COVID-19 situation in India after four contiguous periods of lockdown from March 25 to May 31 (Lockdown 1.0: March 25-April 14^13^, Lockdown 2.0: April 15-May 3^14^, Lockdown 3.0: May 4-May 17^15^, Lockdown 4.0: May 18-May 31^16^). By studying the series of natural experiments across the states and learning from their successes and failures, one has a better likelihood of designing improved targeted interventions for the next phase of the pandemic. Our proposed comprehensive dashboard has broader utility for policymakers and the supporting interactive platform presents daily updates for all metrics and models.

## METHODS

We use publicly available data for all our analyses.^5, 17^ All source code and interactive plots are available at covind19.org.^18^ All computations were done using the RStudio platform.

### Case and Death Counts and Fatality Rates

In addition to simple case and death counts, we look at case-fatality rates (CFR) estimated using all confirmed cases (CFR1, ratio of the total number of deaths and the total number of cases) and closed cases only (CFR2, ratio of the total number of deaths and the sum of the same and the total number of recovered cases). We construct appropriate confidence intervals for these measures.^19^

### Doubling Times and Growth Rates

To quantify the growth of the pandemic, we estimated doubling times (DTs) for total confirmed cases using a 7-day backward-looking window. This measure gives the number of days it would take for total cases to double if its trajectory remained as observed in the past week, and an increase in the DT is evidence of the pandemic slowing down. We also utilize a Bayesian sequential method to estimate the *time-varying effective basic reproduction number*, R, which measures the average number of persons infected by an infected individual. When R falls below 1, the epidemic starts slowing down.^20^

### Testing Summaries

In order to understand the testing landscape, we compute proportion of population tested, test positivity rates (TPRs) and quantify testing shortfalls at the national and state levels. For computing the testing shortfall, we use 2% (roughly the proportion of test-positives in many successful states/regions like Kerala in early phase) as a benchmark. One can change this target based on the stage of the pandemic. The detailed definition of each reported metric and methods for computing corresponding measures of uncertainty are presented in the Supplementary Methods section.

## RESULTS

### Total Number of Cases and Deaths

India had reported its first case of COVID-19 on January 30. The first death from COVID-19 was reported on March 12. In the second week of May, India recorded the highest growth in case counts among Asian countries.^18^ As of June 9, only 4 countries (United States, Brazil, Russia and United Kingdom) had recorded more cases than India.^21^ Supplementary Figure 1 highlights national trends of the COVID-19 outbreak in India by plotting the cumulative number of confirmed cases, fatalities, and recovered cases. To highlight the pronounced geographic pattern across states not visible in Supplementary Figure 1, Figure 1 compares the daily profile of the pandemic at the national level with four states: two that are doing well (Kerala and Punjab) and two that have been hit hard (Maharashtra and Delhi) in terms of the same three compartments. It is clear that Punjab has been doing well and have experienced the first peak, Kerala seems to have many new cases after the strong initial control, Maharashtra has an increasing trend that seems to be stabilizing while Delhi has a high number of cases with a sudden jump in case counts near the end of nation-wide lockdown. Since Maharashtra contributes nearly 35-40% of India’s total number of cases, the national pattern has more resemblance to Maharashtra. Two crucial points emerge from the geographic pattern. First, the concentration of the caseload among the top 10 states has remained relatively stable, at around 90% of the national case count, over this two-month period. Second, the membership of the top 10 states has changed gradually – even as Maharashtra, Delhi and Uttar Pradesh have continued to figure in the list at all four lockdown markers. Supplementary Figures 2 and 3 plot cumulative case and death counts, respectively, across states and over time to highlight these geographic patterns.

**Figure 1.**
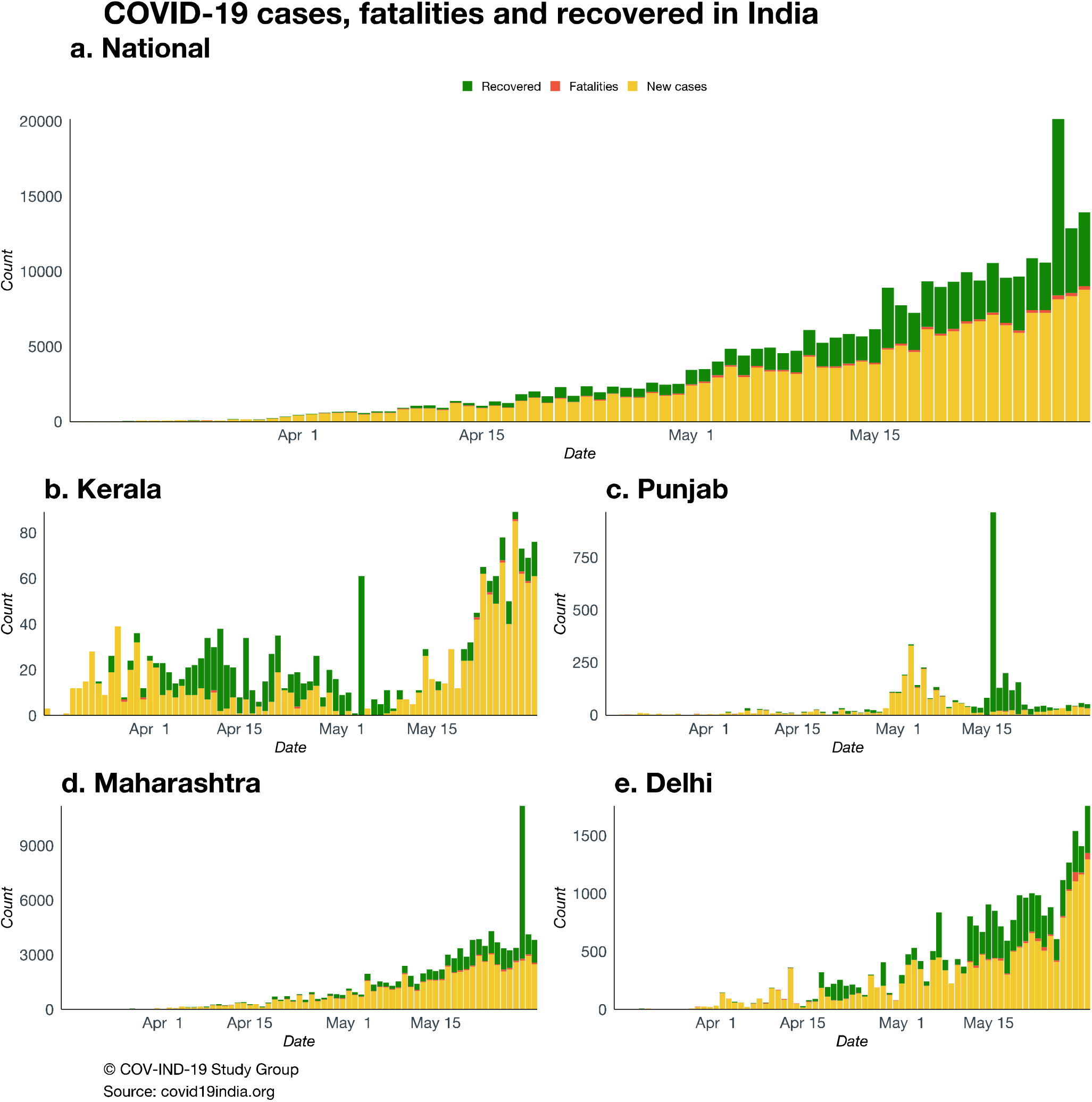
Daily number of reported cases, fatalities, and recovered cases in India over the period between March 15 and May 31 with four states showing the variation. Punjab is an example state of “doing well” whereas case-counts in Delhi and Maharashtra are still increasing. Kerala was doing well initially but has seen a recent surge of cases.

### Case-Fatality Rates

Figure 5a (CFR1) and Supplementary Figure 5 (CFR2) present forest plots of the two estimates of the CFR, along with 95% confidence intervals (CIs), for the 20 states/union territories and for the nation as a whole. To highlight differences across states, we have classified them into three groups: doing well, adequate, needs improvement. The three groups are color-coded red, gray, and green, respectively. Using CFR1, the members of the group that needs improvement (CFR1 >3%) are Gujarat (6.2%), West Bengal (5.8%), Madhya Pradesh (4.3%) Maharashtra (3.4%) and Telangana (3.0%); Haryana (1.0%), Tamil Nadu (0.8%), Kerala (0.8%), Jharkhand (0.8%), Bihar (0.6%), Uttarakhand (0.6%), Odisha (0.5%) and Assam (0.2%) belong to the group that is doing well (CFR1 <1%); all other states fall in the adequate category. Using CFR2, the group needing improvement (CFR2 >5%) now has 6 states/union territories – West Bengal, Gujarat, Maharashtra, Madhya Pradesh, Telangana and Delhi – and the group that is doing well (CFR2 < 3%) has 10 members – Jammu and Kashmir, Andhra Pradesh, Punjab, Jharkhand, Haryana, Kerala, Assam, Bihar, Tamil Nadu and Odisha.

### Doubling Time and Reproduction Number

Figure 2a plots the estimated DTs and Figure 2b plots the estimated time-varying R nationally. Since reliable estimates of DT and R require many days of data, Figures 2a and 2b start on March 15. In both, we report the estimate (along with the 95% CIs for R) on March 24, April 14, May 3, and May 18 corresponding to the initial lockdown and subsequent extensions, in order.

**Figure 2.**
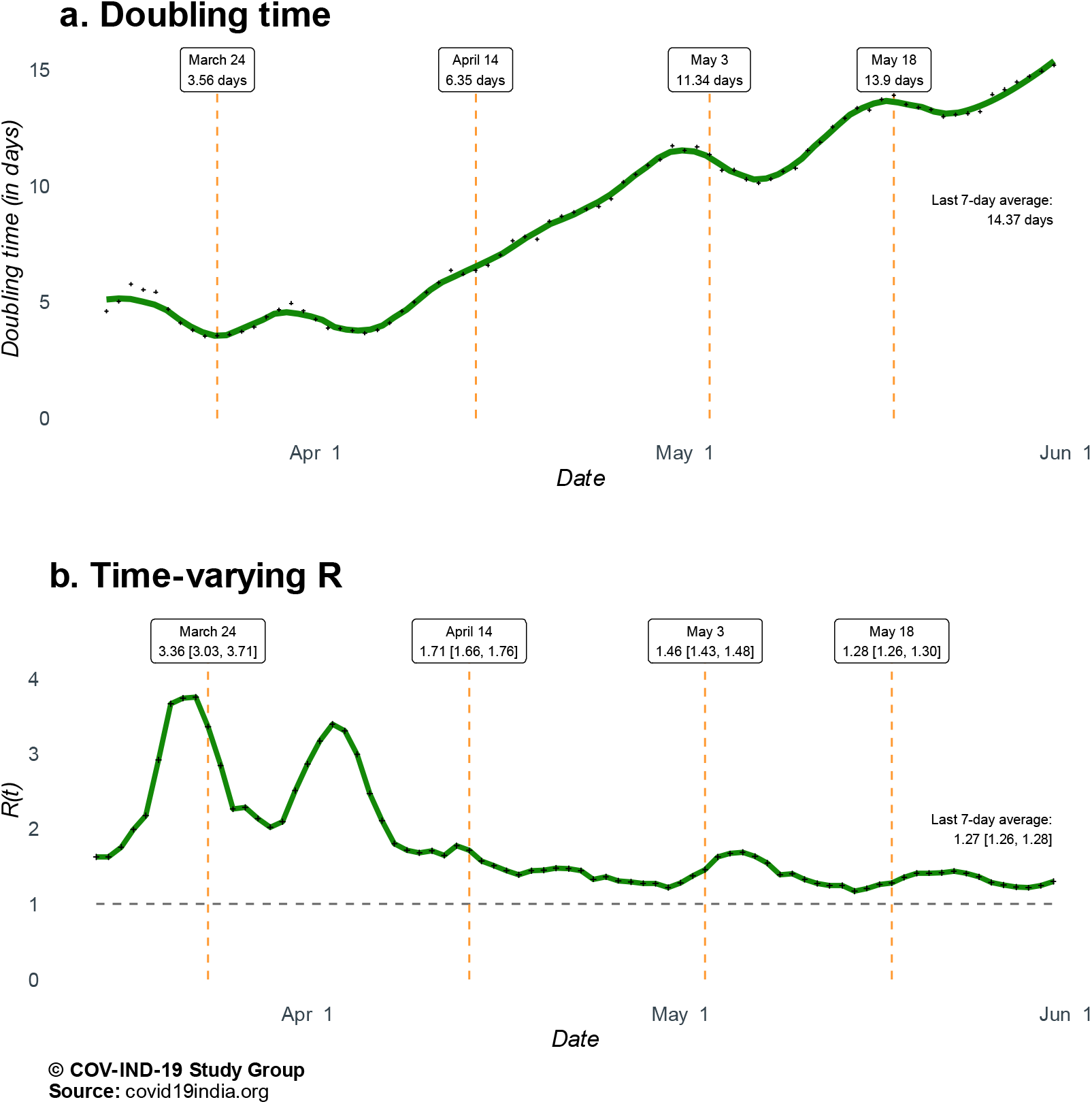
National estimates of doubling times and time-varying R. **Panel a**. Estimated doubling times of total number COVID-19 cases in India, with averages for the pre- and post-lockdown periods and past 7-day average as of May 31. **Panel b**. Estimated time-varying R (effective basic reproduction number) for COVID-19 in India with averages for the pre- and post-lockdown periods and past 7-day average as of May 31, along with 95% confidence intervals.

The time series patterns of estimated DT and R nationally show that the lockdown did slow down the spread of the pandemic. It took about two weeks for the DT to start moving up in a sustained manner. Since early April, the DT has increased from about 5 days to over 14 days by the end of May (Figure 2a). Turning to Figure 2b, we see that the estimated value of R fell over the first lockdown from 3.36 [95% CI: 3.30, 3.71] on March 24 to 1.71 [95% CI: 1.66, 1.76] on April 14, with substantial fluctuation in between. Since then, the estimated R has fallen at a slower pace. The trailing 7-day average value of R for the week ending on May 31 is 1.27 [95% CI: 1.26, 1.28].

These national patterns hide substantial state-level variations, observable in state-level Figures 3a (DT) and 3b (R). Figure 3 a shows that estimated DTs have mostly increased, with Assam, Delhi, Haryana, Odisha, and Uttarakhand being some noteworthy exceptions. Figure 3b indicates that starting from higher values, estimates of R have generally fallen across all states. Again, there are significant differences across states – some states continue to have high values, and in some others, low estimates of R have reverted to relatively high value.

**Figure 3.**
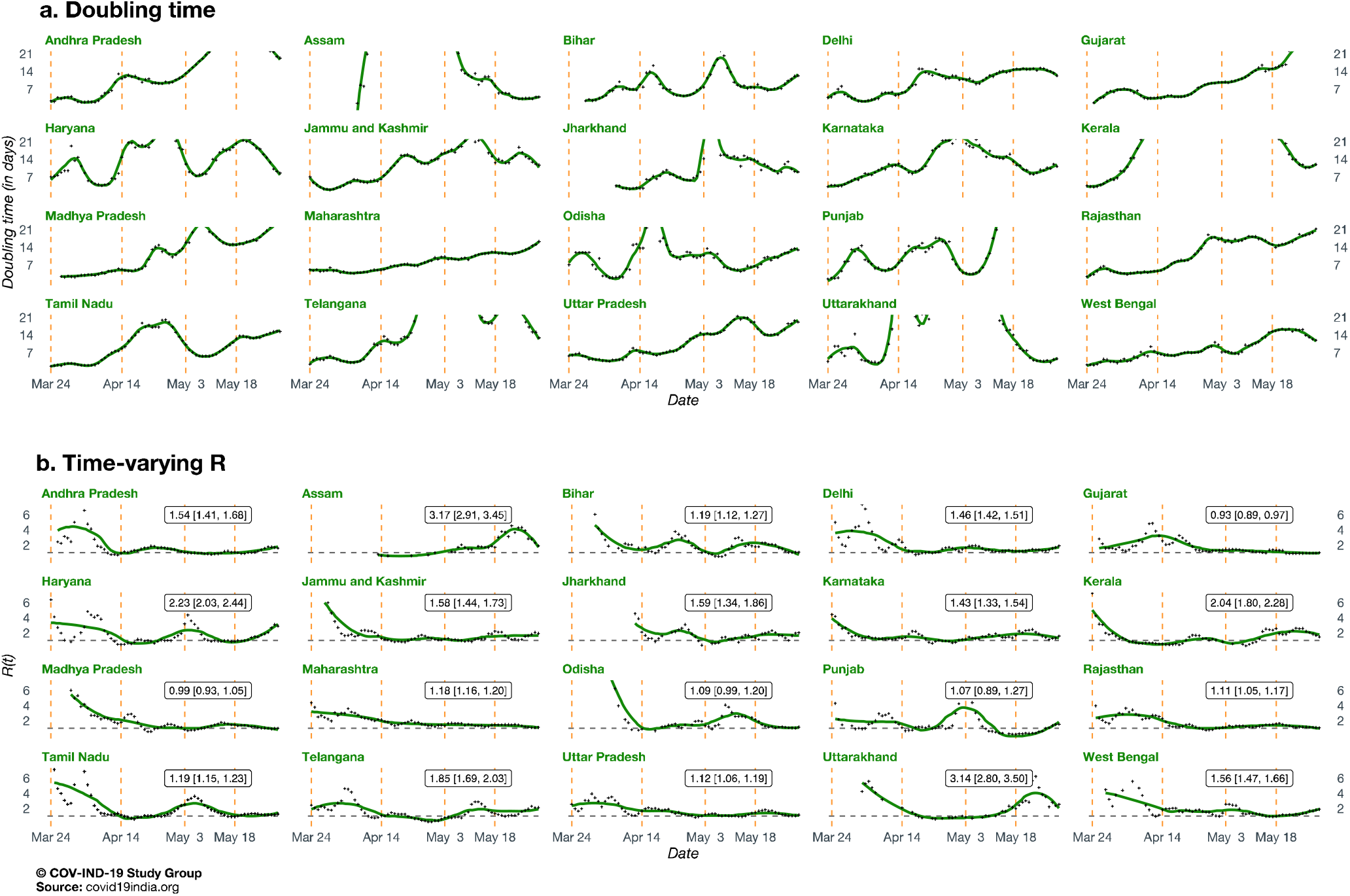
State-wise estimates of doubling times and time-varying R. **Panel a**. Estimated doubling times of total number COVID-19 cases in 20 Indian states and union territories. **Panel b**. Estimated time-varying R (effective basic reproduction number) for COVID-19 in 20 Indian states and union territories along with 95% confidence intervals.

Figure 5b and 5c present forest plots of the average value of the DT (with 7-day range) and estimated R (with 95% CI) over the week before May 31, respectively. Using each figure, we can divide all states into three groups as before. With respect to DT (Figure 5b), the first group (performing well, estimate >two weeks) consists of 11 states. Assam and Uttarakhand are the only 2 states in the third group (needing improvement, estimate <one week). The second group (adequate) consists of 7 states. Similarly, with respect to R (Figure 5c), the well-performing group (estimate <1) consists of only 2 states (Madhya Pradesh and Gujarat). The poor-performing group (estimate >2) consists of 4 states (Assam, Uttarakhand, Haryana and Kerala). All the other states in our sample have average estimated R in [1, 2] (for some states, the CIs include one of the extreme values).

### Testing Coverage and Test Positivity Rate

Going by national-level data, India seems to be doing fairly well in terms of testing. Since mid-April, India’s TPR has fluctuated around 0.04 (Figure 4a). This is lower than not only many European and North American countries, but also significantly lower than its neighbors, like Bangladesh and Pakistan^7, 22^. But this national trend hides the wide variation across states.

**Figure 4.**
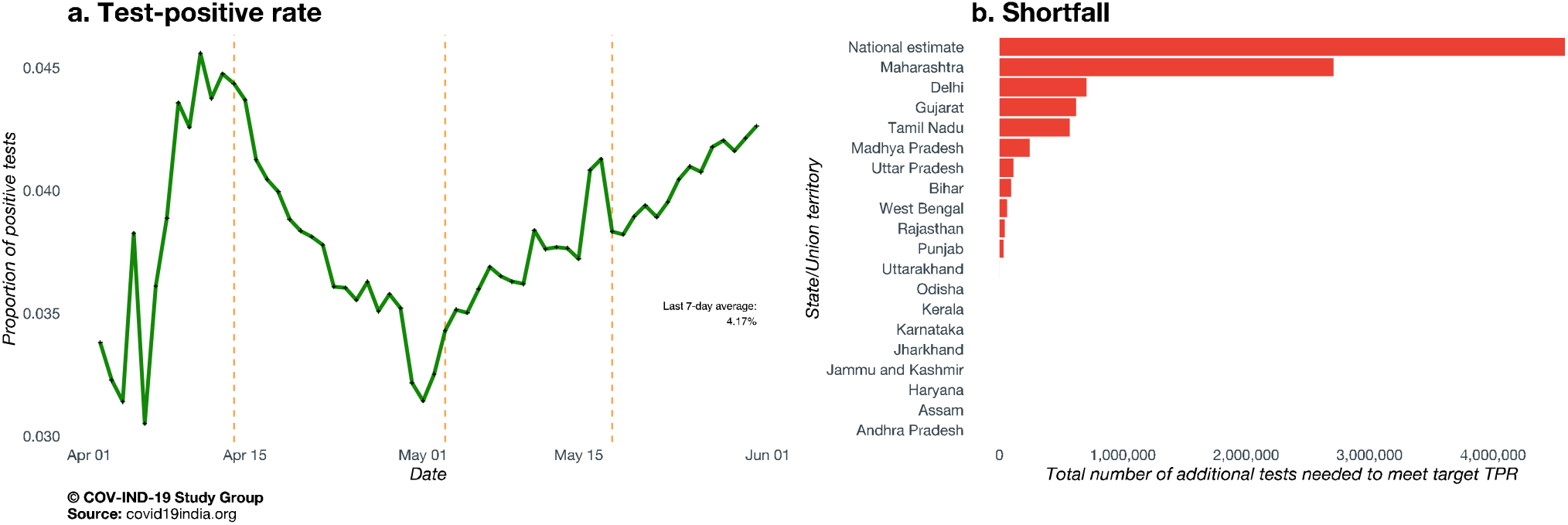
Testing summaries. **Panel a**. Time series plot of test positive rates for India over the period between April 1 and May 31. **Panel b**. Shortfall of number of tests across 20 Indian states and union territories, relative to a benchmark test positive rate of 2%, along with a national estimate. Based on testing data up to May 31.

Supplementary Figure 4 plots the TPR over time for our sample of 20 states/union territories, exhibiting obvious and striking statewide variations. Using a benchmark of 2% TPR to assess adequacy – the rate that some successful states have achieved – we note that many states are far from achieving adequate testing. Even more worrisome is the fact that some states, starting from a value higher than the benchmark, have witnessed rising TPRs. This problem gets compounded since most of these states are where the pandemic is geographically concentrated. Important examples are Delhi, Gujarat, Maharashtra and Tamil Nadu, which have both high case-counts and high/rising TPRs. Bihar, Telangana and Uttarakhand, with relatively low case-counts so far, are witnessing rising TPRs, as seen in Supplementary Table 1 which also contains the proportion of population tested by May 31 across each state.

Figure 4b presents a bar chart of the magnitude of **testing shortfall** across the nation and the 20 states/union territories in our sample. Nationally, we see a shortfall of about 4.58 million tests, with eleven states showing a shortfall, including Maharashtra, Delhi, Gujarat and Tamil Nadu leading the slate. Maharashtra, having done 463,177 tests as of May 31, is the clear outlier in terms of testing shortfall, requiring roughly an additional 2.7 million tests to achieve the adequacy benchmark. For Delhi, Gujarat and Tamil Nadu, the figure is approximately between 560,000 and 700,000 tests. While Madhya Pradesh, Uttar Pradesh, Bihar, West Bengal, Rajasthan, Punjab and Uttarakhand have lower but non-zero testing shortfalls, the other states display adequate testing so far, which definitely is a positive sign.

### Summary State-Level Dashboard: Comprehensive Display of Metrics

With a complete data tsunami, different metrics telling us different features of the pandemic, and a rapidly evolving landscape, we offer a summary dashboard (Figure 5) with classification groups for the states according to various metrics. This captures a snapshot of where things stand across states and the nation, with daily updates available in our app hosted at covind19.org.^18^ To make these data more digestible, Figure 5 follows a consistent color-coding scheme to indicate states/union territories that are doing well (green) and states/union territories needing improvement (red).

**Figure 5.**
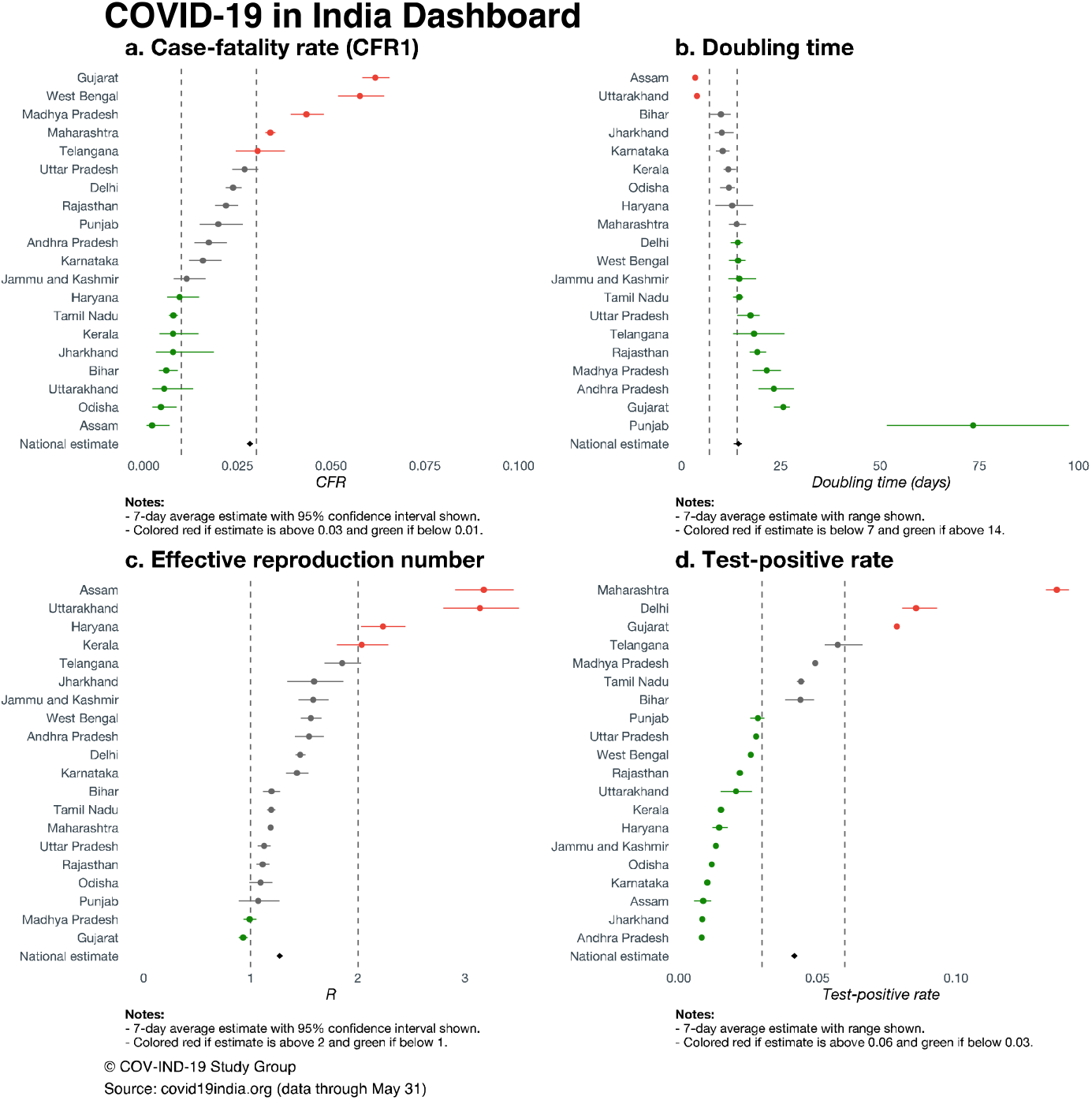
Forest plot dashboard. **Panel a**. Forest plot of estimated case-fatality rates (CFR1) based on all confirmed cases as of May 31, along with 95% confidence intervals, for 20 states and union territories of India, and a national summary. **Panel b**. Forest plot of estimated doubling times (in days) based on data from a 7-day past window from May 31, along with 95% confidence intervals, for 20 states and union territories of India, and a national summary. **Panel c**. Forest plot of estimated time-varying R (effective basic reproduction number) based on data from a 7-day past window from May 31, along with 95% confidence intervals, for 20 states and union territories of India, and a national summary. **Panel d**. Forest plot of test positive rates (proportion scale) based on data as of May 31, for 20 states and union territories of India, along with a national summary.

Panel a shows CFR1 along with the 95% CI (doing well: below 1%; needs improvement: above 3%). While the all-India CFR1 on May 31 was 2.84%, state-level CFR1s ranged from 6.2% (Gujarat) to 0.2% (Assam). Panel b shows the 7-day average DT along with the range (doing well: above 14 days; needs improvement: below 7 days). The quickest DT is in Assam (3.5 days, range: [3.1, 4.0]), while the slowest DT is in Punjab (73.5 days, range: [51.6, 97.5]). The national estimate is 14.4 days (range: [13.2, 15.2]), with about half of states having DT exceeding 14 days.

Panel c shows the 7-day average R along with the 95% CIs (doing well: below 1; needs improvement: above 2). We see that 7-day average estimates range from 0.93 (95% CI: [0.88, 0.97]) in Gujarat to 3.17 (95% CI: [2.91, 3.45]) in Assam, with a national estimate of 1.27 (95% CI: [1.26, 1.28]). Panels b and c exhibit how the DT, a function of cumulative cases, is less sensitive to daily movements than R. For example, Kerala has done well controlling the outbreak in terms of DT, but a small recent increase in observed cases results in a 7-day average R estimate close to 2.

Panel d shows the 7-day average TPR along with the range (doing well: below 3%; needs improvement: above 6%). The lowest 7-day average TPR is seen in Andhra Pradesh (0.83%, range: [0.81%, 0.85%]), with the highest being seen in Maharashtra (13.63%, range: [13.25%, 14.07%]). Generally, states with larger cumulative case counts are seen to have higher TPRs. Considering the test shortfall (Figure 4b), this is likely explained by inadequate number of tests being performed given the scale of the outbreak in these places. The national 7-day average TPR is 4.17% (range: [4.08%, 4.26%]).

It is important to consider these metrics together, keeping their nuances in mind:

- CFR1 is an indicator of the fatality associated with the epidemic, but its value is sensitive to the number of tests being performed. A high CFR1 might very well arise from inadequate testing. Hence, the CFR1 is best used in conjunction with some measure of adequate testing.
- R can indicate a recent outbreak but is sensitive to the level of daily cases being observed (i.e., a state/union territory with few cases can have a high R). In parallel, DT is a longer-term measure since it is a function of cumulative cases (i.e., this metric is more robust to fluctuations in recent daily cases). These are relative metrics and do not inform us about projected health care needs.
- TPR is both a function of the size of the outbreak in an area and the number of tests being performed. A higher TPR can indicate insufficient levels of testing and selective testing of symptomatic patients. The proposed metric of testing shortfall is a policy relevant metric.

## DISCUSSION

While it is common for analysts and policymakers to predict a peak for the COVID-19 in India^23^, our analysis shows that the concept of a peak for the whole country is, at best, ambiguous^24^. Differences in estimates of R (Figure 3b) and estimated DTs (Figure 3a) suggest that peaks will vary across states. Predictions from the extended SIR (eSIR)^25^ model available at covind19.org show that peak in case-counts might start as early as the end of July in some states and go all the way to September in many others. Some states like Punjab have already experienced their first peak. These predictions are in line with basic intuition about the dynamics of the pandemic in India. Initial cases were imported, and the initial growth was limited to a few states which saw the arrival of international travelers. These initial cases seeded the epidemic and saw the explosion of cases. With the non-medical intervention of lockdown, mobility was limited at the macro level (inter-state, inter-city), which reduced transmission rates (Figures 2-3). Pre-lockdown infections and micro-mobility resulted in growth of cases within states; notably, the top 10 states on April 18 and May 31 are largely same. Now that we are in the targeted lockdown phase, internal migration will start playing an increasingly important role in the spread of the pandemic.

India has a large migrant worker population. Estimates of out-of-state and out-of-district migrants ranged from 60 million to 80 million in 2011, and average work-related migration flows between states over the period 2011-16 was about 9 million per year.^26^ With the easing of lockdown and work slowly resuming, a large migrant population will soon start traveling back to their workplaces and India could see the next surge in cases in states home to higher numbers of migrant workers. A combined and rigorous strategy of testing suspected patients, tracing contacts of patients, and isolating infected persons can effectively break the chain of transmission and slow down the pandemic. In a country like India, which can ill afford the severe economic disruption caused by a lockdown, this alternative approach has much to recommend itself.^27^

While looking at testing coverage is a common way to gauge adequacy of testing rates, an alternative approach is to track the TPR, i.e., fraction of positives in the total number of persons tested.^28^ High and/or rising TPRs indicate that only those with a high probability of having the SARS-COV-2 are being tested. These would mostly be patients with strong symptoms of the disease. Hence, many patients with lower probability of being infected and many yet to show symptoms would be out of the ambit of testing. Thus, the estimated prevalence of the disease would be a gross underestimate of the ‘true’ prevalence rate.

Given the spatial and temporal pattern of the pandemic’s spread, it is extremely important to prioritize policies. Resources must be mobilized to help one cluster of states and then move to the next cluster. It might be useful for the central government and the Indian Council of Medical Research (ICMR) to classify states in terms of the phases of the epidemic. Even as the worst-hit states are being addressed, the next set could be put on high alert. It is this dynamic policy intervention that will be required to deal effectively with the cascading pattern of the pandemic across Indian states.

In implementing such a dynamic policy, it is extremely important to facilitate replication of successful strategies across states. Kerala’s rapid response in terms of testing, contact tracing and quarantining; Odisha and Kerala’s use of local governance structures and community health networks for surveillance and dissemination of correct information; Punjab’s use of data analytics and district level granular contact tracing, tracking and isolation – all these experiences will be of use in other states that are likely to see a surge in cases in the coming weeks.

The success of some states gives us hope that there are strategies to beat the virus that have worked well. Resources can be mobilized and optimally deployed to address the acute situations in high density population areas like Maharashtra, Gujarat and Delhi. In all these efforts, nuanced state-level summaries offer their utility.

## Data Availability

All source code and interactive plots are available at covind19.org.

http://covind19.org/

